# Association of *IFNAR2* rs2236757 and *OAS3* rs10735079 polymorphisms with susceptibility to COVID-19 infection and severity

**DOI:** 10.1101/2022.09.27.22280425

**Authors:** Mohammad Abdelhafez, Abedelmajeed Nasereddin, Omar Abu Shamma, Rajaa Abed, Raghida Sinnokrot, Omar Marof, Tariq Heif, Zaid Erekat, Amer Al-Jawabreh, Suheir Ereqat

**Affiliations:** Faculty of Medicine, Al-Quds University, Abu Deis, East Jerusalem, Palestine; Biochemistry and Molecular Biology Department, Faculty of Medicine, Al-Quds University, Abu Deis, East Jerusalem, Palestine; Department of Medical Laboratory Sciences, Faculty of Allied Health Sciences, Arab American University, Jenin, Palestine

**Keywords:** SARS-CoV-2, COVID-19, SNPs, *OAS3*, *IFNAR2*

## Abstract

The clinical course and severity of COVID-19 vary among patients. This study aimed to investigate the association of the interferon receptor (*IFNAR2*) rs2236757 and oligoadenylate synthetase 3 (*OAS3*) rs10735079 gene polymorphisms with risk of COVID-19 infection and severity among Palestinian patients. The study was conducted between April and May 2021 on 154 participants that divided into three groups: the control group (RT-PCR-negative, n=52), the community cases group (RT-PCR-positive, n= 70) and the critically ill cases (ICU group; n=32). Genotyping of the studied polymorphisms was conducted by amplicon-based next-generation sequencing.

The genotype distribution of the *IFNAR2* rs2236757 was significantly different among the study groups (P = 0.001), while no significant differences were observed in the distribution of *OAS3* rs10735079 genotypes (P = 0.091). Logistic regression analysis adjusted for possible confounding factors revealed a significant association between the risk allele rs2236757A and critical COVID-19 illness (P < 0.025). Among all patients, the rs2236757GA carriers were more likely to have sore throat (OR, 2.52 (95% CI 1.02-6.24); P = 0.011); the risk allele rs2236757A was associated with dyspnea (OR, 4.70 (95% CI 1.80-12.27); P < 0.001), while the rs10735079A carriers were less prone to develop muscle aches (OR, 0.34 (95% CI 0.13-0.88); P = 0.0248) and sore throat (OR, 0.17 (95% CI 0.05-0.55); P < 0.001). In conclusion, our results revealed that the rs2236757A variant was associated with critical COVID-19 illness and dyspnea, whereas the rs10735079A variant was protective for muscle aches and sore throat.

## Introduction

The Coronavirus disease 2019 (COVID-19) is a respiratory and systemic disease caused by severe acute respiratory syndrome coronavirus 2 (SARS-CoV-2), first reported in Wuhan, China, in December 2019 and then rapidly spread across the globe. Worldwide, more than 613 million coronavirus patient was reported with more than 6.5 million of total deaths (Worldometers.info: Dover, Delaware, U.S.A) accessed date 10-09-2022). It is transmitted predominately from person to person mainly through inhalation of small, exhaled respiratory droplets containing infectious virions [1]. The clinical manifestations range from asymptomatic, to severe, life-threatening acute respiratory distress syndrome (ARDS), multi-organ failure, and death [2]. The major symptoms of the disease include fever, cough, fatigue, sore throat, headache, and shortness of breath, with progression to pneumonia [3]. Households are favorable venues for viral transmission, where family members may crowd and be in close contact without following social distancing rules or using masks. Although all household contacts of a positive COVID-19 patient are exposed to the virus, not all get infected.

Previous reports showed that male gender, elder age group, and the existence of comorbidities (e.g., cardiovascular, pulmonary, and renal diseases) are risk factors for COVID-19 infection [4]. However, the wide range of the reported symptoms suggested that genetic risk factors may also have a crucial role in disease progression. Although the virus’s new mutations have emerged (e.g., UK, South Africa, and India), few studies described the inter-individual genetic differences in the immune response to these new versions of coronavirus.

It was reported that variants of the angiotensin converting enzyme 2 (ACE2) gene encode the cellular receptor for SARS-CoV-2 and polymorphisms of serine protease TMPRSS2 affect viral entry and invasion and thus increase COVID-19 severity [5,6]. Moreover, a genome-wide association study (GWAS) conducted in the UK compared the genetic variants in critically ill patients (n= 2244) with severe COVID-19 to variants found in a healthy control group. The study revealed significant associations between the severity of COVID-19 and the genetic variants in five loci including chromosome 3p21.31, spanning the *SLC6A20, LZTFL1, CCR9, FYCO1, CXCR6*, and *XCR1* genes; chromosome 12q24.13, in the *OAS* gene cluster; chromosome 19p13.2, near tyrosine kinase 2 (TYK2); chromosome 19p13.3, within dipeptidyl peptidase 9 (*DPP9*); chromosome 21q22.1, within the interferon receptor gene *INFAR2*. Of these genes, *IFNAR2* and *OAS*, are important in the early stages of the disease, whereas the *DPP9, TYK2*, and *CCR2* genes drive inflammatory processes in the late stages of critical COVID-19 [7].

. It is well-established that SARS-CoV-2 infection activates innate and adaptive immune response, failure of this system and dysregulated massive pro-inflammatory host response would cause harmful tissue damage [8].

In this context, the study objective was to investigate the association of four SNPs (*IFNAR2* rs2236757, *DPP9* rs2109069, *OAS3* rs10735079, and *LZTFL1* rs73064425 variants), with the susceptibility to COVID-19 infection in Palestinian household contacts and their associations with the clinical manifestations and severity of COVID 19, using amplicon-based next generation sequencing (NGS).

## Materials and methods Study participants

The study participants were recruited from 30 household Palestinian families-living in different cities in the West Bank, Palestine-between April and May 2021. The family was included in the study if it has at least one clinically and laboratory-confirmed COVID-19 case and one laboratory-negative household contact with no COVID-19 symptoms. We divided all family members into two groups: infected cases (community cases group) or uninfected household contacts (control group). An infected case was defined by having a positive RT-PCR test regardless of having symptoms or not and regardless of being a primary or secondary case. An uninfected contact was defined as a family member who had unprotected contact with a positive case, lives at the same place, stayed asymptomatic for ten days after symptoms onset or RT-PCR diagnosis of a positive case; and was tested negative by RT-PCR. Additionally, we consecutively enrolled patients from an intensive care unit (ICU) -at the Palestinian Medical Complex, Ramallah - due to critical COVID-19 illness (ICU cases).

We excluded individuals who received COVID-19 vaccination, regardless of the type of the vaccine. Patients’ data, including demographic information, symptoms, RT-PCR test result, and comorbidities, were collected via a well-structured questionnaire supervised by health care personnel.

### Sampling, DNA extraction, and genotyping

Blood-sample (five ml) was collected in an EDTA tube from all study participants. The DNA was extracted from each blood sample (200 μl) using a genomic QIAamp DNA purification kit as per the manufacturer’s instructions (Qiagen, Hilden, Germany) and kept frozen (−20C) for further analysis. All DNA samples were genotyped for the rs2236757 of *IFNAR2*, rs2109069 of *DPP9*, rs10735079 of *OAS3*, and rs73064425 of the *LZTFL1* gene using amplicon-based NGS (NGS). Briefly, two primers (forward and reverse) were used to target each single nucleotide polymorphism (SNP) as described in Table S1. All primers were modified with over-hanged Illumina adaptor sequences at the 5′ ends (bolded, Table S1), targeting partial sequences of the studied genes. The final product size for each targeted gene was mentioned in Table S1.

The PCR product was visualized by 1.5% agarose gel, cleaned by Agencourt AMPure XP system (X1, A63881; Beckman Coulter Genomics, Indianapolis, IN, USA), and eluted into a final volume of 25 μl. All purified samples were amplified by dual indices PCR to barcode each sample using Nextera XT Index Kit (Illumina, San Diego, CA, USA); Five μl from each barcoded sample were pooled together, cleaned again by Agencourt AMPure XP system (X1), and eluted in 50 μl elution buffer. The concentrations of the prepared Libraries were tested by Qubit® Fluorometer (Invitrogen, Carlsbad, CA, USA). A concentration of 4 nM was used with a target of 20k reads for each sample. Deep sequencing was done by NextSeq 500/550 machine using 150-cycle Mid Output Kit (Illumina, San Diego, CA, USA).

### Bioinformatics and Sequence Analysis

The sequencing data were uploaded to the Galaxy web platform, and we used the public server at “usegalaxy.org” to analyze the obtained DNA sequences [9]. The workflow of filtration included Illumina adaptor trim and quality selection of Q > 20 with a minimal read length of 100 bp. We used eight virtual probe sequences to identify the targeted variants (Table S1). Ultimately, the genotypes were determined based on the ratio between the read counts for wild-type and minor alleles. SNPs were included in the study if they passed our quality measures; Hardy–Weinberg equilibrium (HWE) > 0.05 and genotyping rate > 95%.

### Statistical Analysis

We performed the statistical analysis using the SPSS package, version 26.0 (SPSS, Inc., Chicago, IL, USA) and the R environment v.4.1.3. All tests were two-tailed, and we considered P-value < 0.05 significant unless specified. We tested for The Hardy-Weinberg equilibrium (HWE) for all SNPs using the “SNPassoc” package [10]. Moreover, we examined the genetic susceptibility of SARS-CoV-2 and the genetic association with the critical COVID-19 illness by comparing the community patients with the controls, and the ICU patients with the controls, respectively, using five genetic models (co-dominant, dominant, over-dominant, recessive, and additive). Models were adjusted for patient characteristics; age, gender, smoking, history, hypertension, diabetes mellitus, and coronary artery diseases using the “SNPassoc” package. Adjusted odds ratios (ORs) with the associated 95% confidence interval (CI) were calculated for each model. The same models were used to investigate Gene-(Symptoms/Signs) associations. The best model for each SNP was selected using the Akaike information criterion [11].We used Bonferroni correction for multiple comparisons to correct statistical significance (P < 0.05, divided by the number of analyzed SNPs) [12,13]. Ultimately, we investigated for any potential gene-gene interaction.

## Results

### Characteristics of Study Participants

A total of 154 Palestinians were included in this study and divided into three groups; COVID-19 infected patients (Community cases group, n=70), uninfected household contacts (Control group, n=52) and critically ill COVID-19 patients (ICU group, n=32). In each group, the median (IQR) age was 28 (27), 24.5 (21.5), and 61 years (27). The characteristics and comorbidities of each study group are shown in Table 1. The median age, the prevalence of smoking, diabetes mellitus (DM), hypertension, and coronary artery disease (CAD) were significantly higher in the ICU group (P < 0.05). The clinical characteristics of COVID-19 patients in the community and ICU groups with signs and symptoms frequencies are presented in Table 2. The percentage of symptomatic patients was 93% in the community cases group, whereas 100% in the ICU group. The frequency of fatigability, headache, and loss of taste and/ or smell was significantly higher in the community cases group (P < 0.05). However, dyspnea and cough were more frequent in the ICU group (P < 0.05).

**Table 1:** Baseline characteristics and comorbidities.

|  | Community patients<br>(N=70) | ICU patients<br>(N=32) | Control (N=52) | P- Value |
| --- | --- | --- | --- | --- |
| **M: F** | 32:38 | 19:13 | 33:19 | 0.124 |
| **Age †** | 28 [21.5- 48.5] | 61 [41-68] | 24.5 [20.25- 41.75] | **<0.001\*** |
| **Smoking** | 15.70% | 31.30% | 30.80% | 0.117 |
| **DM** | 5.70% | 25% | 3.80% | **0.009** |
| **Hypertension** | 4.30% | 43.80% | 3.80% | **0.001** |
| **CAD** | 4.30% | 40.60% | 0% | **0.001** |

† Median [25<sup>th</sup> -75<sup>th</sup> percentile]. \* P-value was obtained by Independent-Samples Kruskal-Wallis Test. P < 0.05 was considered significant. M, male; F, female; DM, diabetes mellitus; CAD, coronary artery disease; ICU, intensive care unit.

**Table 2:** Signs and symptoms of COVID-19 in the community cases and the intensive care unit groups

|  | Community cases<br>group(70) |  | ICU group(32) |  | P-value |
| --- | --- | --- | --- | --- | --- |
|  | N | % | N | % |  |
| **Symptomatic** | 64 | 93 | 32 | 100 | 0.118 |
| **Runny nose** | 34 | 49 | 7 | 22 | **0.009** |
| **Fatigue** | 52 | 75 | 11 | 36 | **0.001** |
| **Headache** | 49 | 71 | 10 | 33 | **0.001** |
| **Fever** | 43 | 62 | 21 | 70 | 0.463 |
| **Loss of smell/ taste** | 45 | 65 | 12 | 40 | **0.02** |
| **Muscle ache** | 42 | 61 | 21 | 68 | 0.51 |
| **Diarrhea** | 24 | 35 | 6 | 19 | 0.119 |
| **Sore throat** | 27 | 39 | 9 | 30 | 0.385 |
| **Cough** | 33 | 49 | 22 | 71 | 0.037 |
| **Dyspnea** | 24 | 35 | 27 | 87 | **0.001** |

P < 0.05 was considered significant. ICU, intensive care unit.

### Genotyping of *IFNAR2* rs2236757 and *OAS3* rs10735079

The minor allele frequency (MAF) of the *IFNAR2* rs2236757A and the *OAS3* rs10735079A was 27% and 50%, respectively. The frequency and genotypes distribution of the *IFNAR2* rs2236757A and the *OAS3* rs10735079A among the three study groups are provided in Table 3. The *IFNAR2* rs2236757 genotypes distribution was significantly variable among the study groups (P = 0.001), while no significant differences were observed in the distribution of *OAS3* rs10735079 genotypes (P = 0.091). The two SNPs; *DPP9* rs2109069, and the *LZTFL1* rs73064425, were excluded from the study due to deviation from the HWE (i.e., P < 0.05) and the low genotyping rate (i.e., <95%).

**Table 3:** Genotypes distribution of the *IFNAR2* rs2236757 and *OAS3* rs10735079 among the studied groups

| SNP | Alleles (major/minor) | Minor allele frequency N (%) | Genotypes | Community cases group | ICU group | Control group | P- value |
| --- | --- | --- | --- | --- | --- | --- | --- |
| rs2236757 (*IFNAR2*) | G/A | 83 (26.9) | AA | 0 | 6 | 2 | 0.001 |
|  |  |  | GA | 35 | 15 | 17 |  |
|  |  |  | GG | 35 | 11 | 33 |  |
| rs10735079 (*OAS3*) | G/A | 153 (50.3) | AA | 20 | 11 | 12 | 0.091 |
|  |  |  | GA | 31 | 8 | 28 |  |
|  |  |  | GG | 19 | 13 | 10 |  |

Person's chi-squared test, P < 0.05 was considered significant.

### *IFNAR2* rs2236757 and *OAS3* rs10735079 polymorphisms and susceptibility to COVID-19 infection

Logistic regression analysis under five genetic models adjusted for age, sex, smoking history, DM, hypertension, and CAD was used to investigate the role of *IFNAR2* rs2236757 and *OAS3* rs10735079 polymorphisms in the susceptibility of COVID-19 infection and severity, the community cases group was compared to the control group and the ICU group was compared to the control group. As shown in Table 4, none of the studied polymorphisms have a statistically significant association with SARS-CoV-2 infection among community cases (P > 0.025) after Bonferroni correction. However, the risk allele rs2236757A of the *IFNAR2* gene was significantly associated with critical COVID-19 illness in all genetic models (P < 0.025) except for the recessive (P = 0.4). According to the Akaike information criterion, the dominant model was the best to explain the association (OR, 8.65 (95% CI 1.60-46.68); P = 0.005). No significant relationship between the *OAS3* rs10735079 polymorphism and critical COVID-19 illness was observed (P > 0.025; Table S2).

**Table 4:**
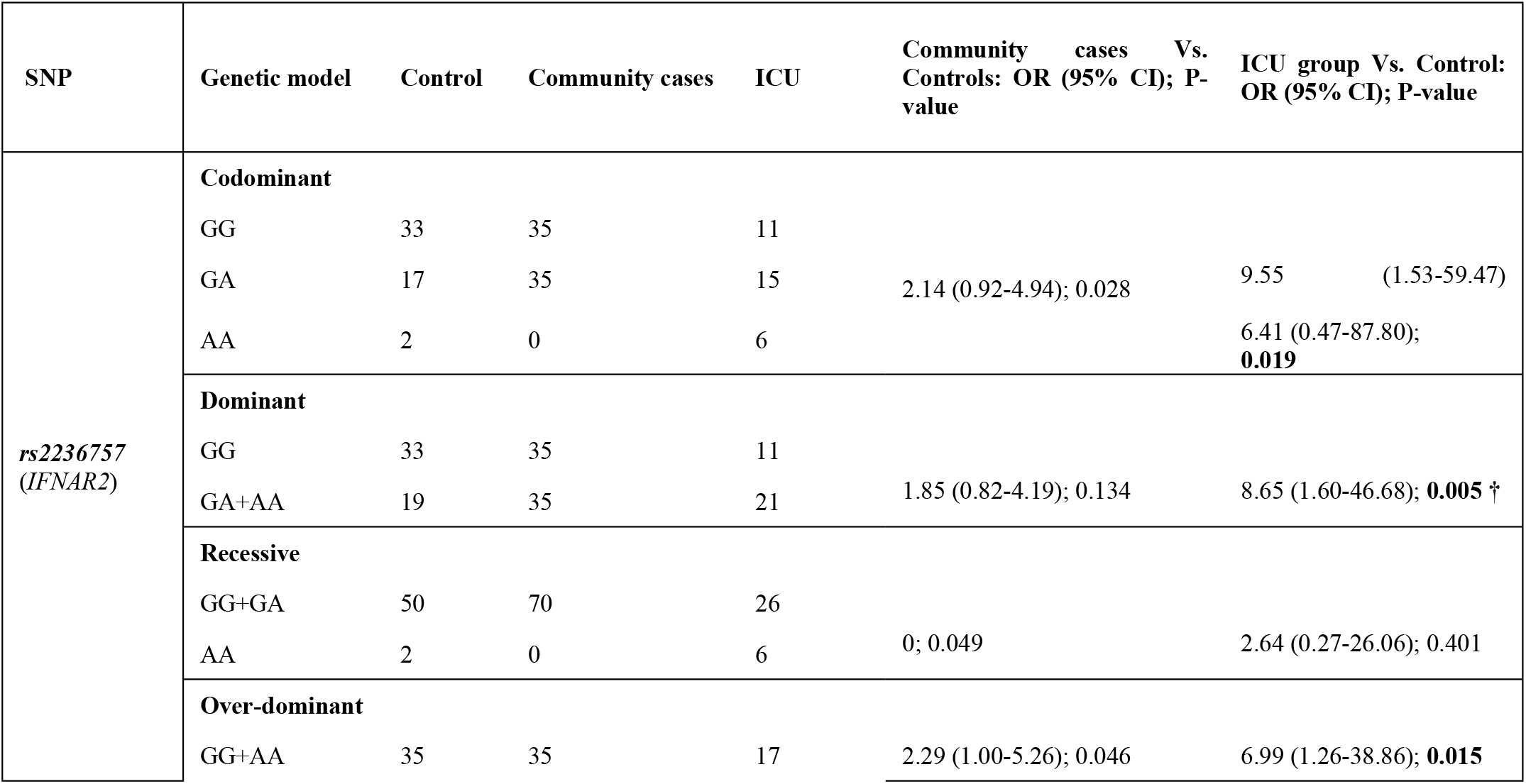
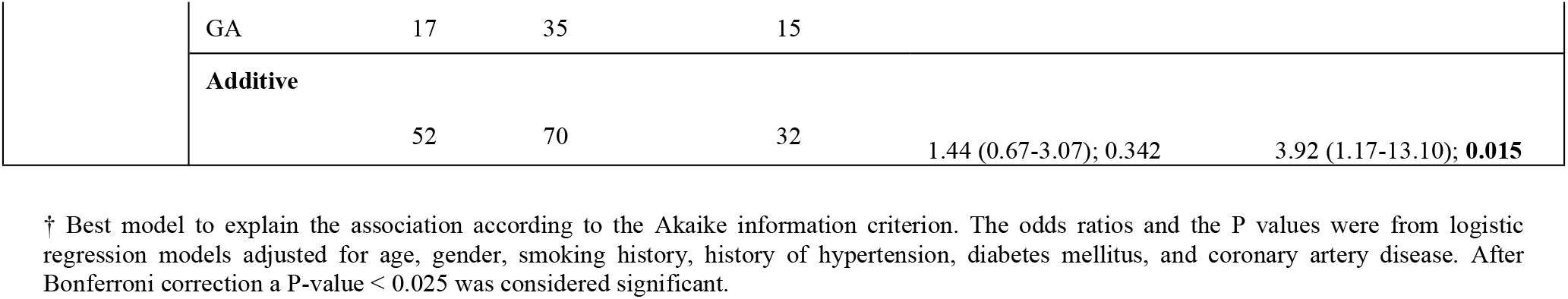
Association of *IFNAR2* rs2236757 polymorphism with COVID-19 infection among the studied groups

| SNP | Genetic model | Control | Community cases | ICU | Community cases Vs. Controls: OR (95% CI); P-value | ICU group Vs. Control: OR (95% CI); P-value |
| --- | --- | --- | --- | --- | --- | --- |
| *rs2236757* (*IFNAR2*) | **Codominant** |  |  |  |  |  |
|  | GG | 33 | 35 | 11 |  |  |
|  | GA | 17 | 35 | 15 | 2.14 (0.92-4.94); 0.028 | 9.55 (1.53-59.47) |
|  | AA | 2 | 0 | 6 |  | 6.41 (0.47-87.80); **0.019** |
|  | **Dominant** |  |  |  |  |  |
|  | GG | 33 | 35 | 11 |  |  |
|  | GA+AA | 19 | 35 | 21 | 1.85 (0.82-4.19); 0.134 | 8.65 (1.60-46.68); **0.005** † |
|  | **Recessive** |  |  |  |  |  |
|  | GG+GA | 50 | 70 | 26 |  |  |
|  | AA | 2 | 0 | 6 | 0; 0.049 | 2.64 (0.27-26.06); 0.401 |
|  | **Over-dominant** |  |  |  |  |  |
|  | GG+AA | 35 | 35 | 17 | 2.29 (1.00-5.26); 0.046 | 6.99 (1.26-38.86); **0.015** |
|  | GA | 17 | 35 | 15 |  |  |
|  | **Additive** |  |  |  |  |  |
|  |  | 52 | 70 | 32 | 1.44 (0.67-3.07); 0.342 | 3.92 (1.17-13.10); **0.015** |

† Best model to explain the association according to the Akaike information criterion. The odds ratios and the P values were from logistic regression models adjusted for age, gender, smoking history, history of hypertension, diabetes mellitus, and coronary artery disease. After Bonferroni correction a P-value < 0.025 was considered significant.

### Association of *IFNAR2* rs2236757 and *OAS3* rs10735079 polymorphisms with COVID-19 signs and symptoms

Logistic regression analysis under five genetic models adjusted for age, sex, smoking history, DM, hypertension, and CAD was used to investigate the association of *IFNAR2* rs2236757 and *OAS3* rs10735079 polymorphisms with COVID-19 signs and symptoms. For all patients (the community cases group and ICU group), the *IFNAR2* rs2236757 GA carriers were more likely to have a sore throat (OR, 2.52 (95% CI 951.02-6.24); P = 0.011) (Table 5). In addition, patients who developed dyspnea were more likely to have the risk allele rs2236757A; the association was best explained by the additive model (OR, 4.70 (95% CI 1.80-12.27); P < 0.001). On the other hand, patients with the risk allele rs10735079A were less prone to develop muscle aches (OR, 0.34 (95% CI 0.13-0.88); P = 0.0248) and sore throat (OR, 0.17 (95% CI 0.05-0.55); P < 0.001), both associations were best explained by the recessive model (Table 5). Further analysis was performed to compare the community cases group with the control group and the ICU group with the control group as shown in Table 6. None of the community cases group was homozygous for the risk allele *IFNAR2* rs2236757A (Table 6). However, the risk allele rs2236757A was associated with loss of taste or smell (OR, 3.57 (95% CI 1.19-10.72); P = 0.019), muscle aches (OR, 3.65 (95% CI 1.12-11.86); P = 0.025), and dyspnea (OR, 4.84 (95% CI 1.45-16.13); P = 0.006). We also found that patients with sore throat in the community cases group were unlikely to be homozygous (AA) for the risk allele rs10735079A of *OAS3*; the association was only explained by the recessive model (OR, 0.19 (95% CI 0.05-0.79); P = 0.012). Among the ICU group, muscle aches were the only symptoms that had a genetic association; patients with the risk allele rs10735079A were less prone to muscle aches in two models (recessive and additive), and best explained by the additive (OR, 0.22 (0.05-0.99); P = 0.014) (Table 6). We tested gene-gene interaction in all models that had a significant association with the clinical manifestations; we did not find a statistically significant interaction between the SNPs. Still, heterozygous rs2236757 (GA) carriers with sore throat were less likely to be homozygous (AA) for the risk allele rs10735079A (OR, 0.06 (95% CI 0.01-0.57)) (data not shown).

**Table 5:**
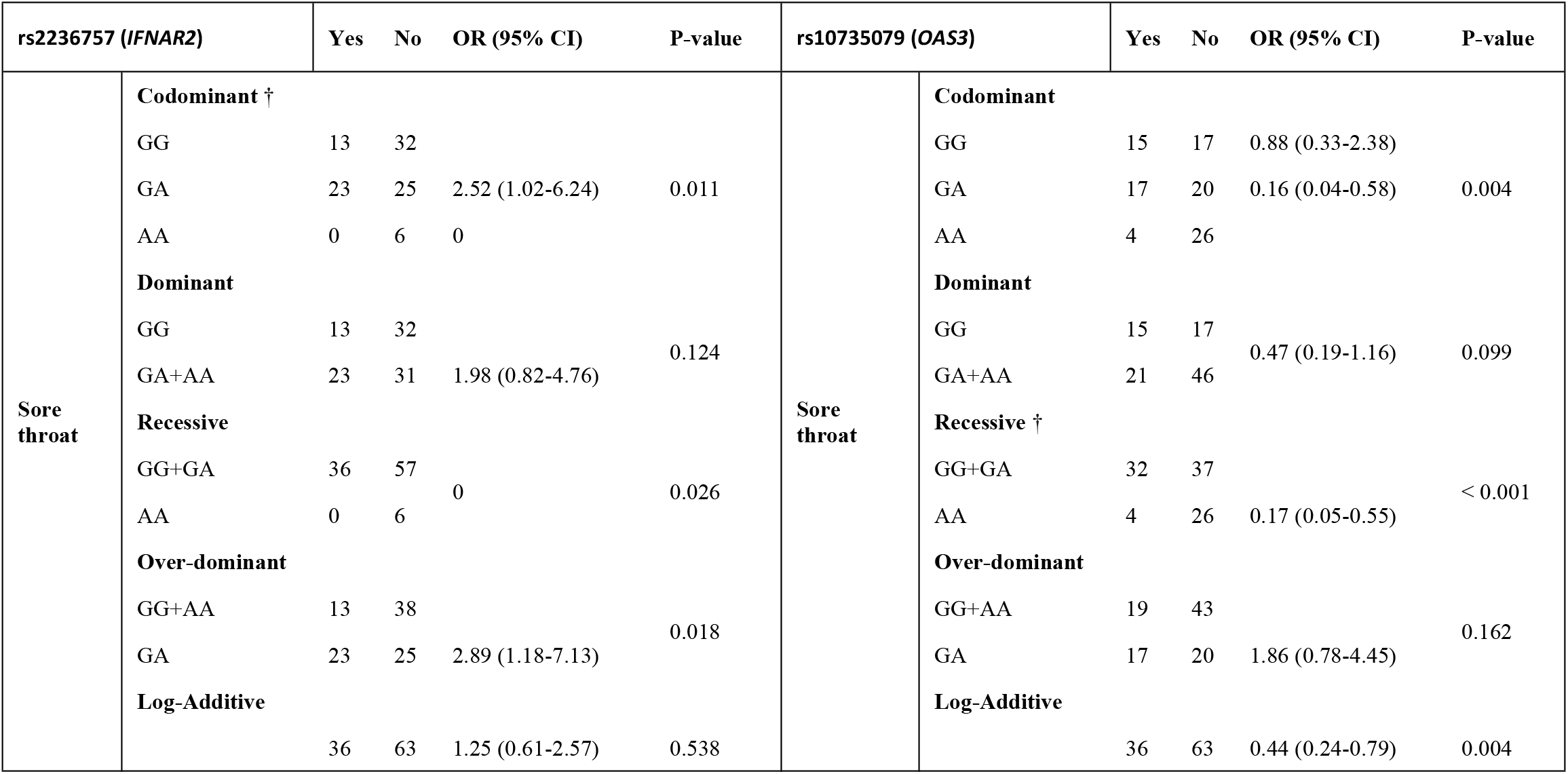
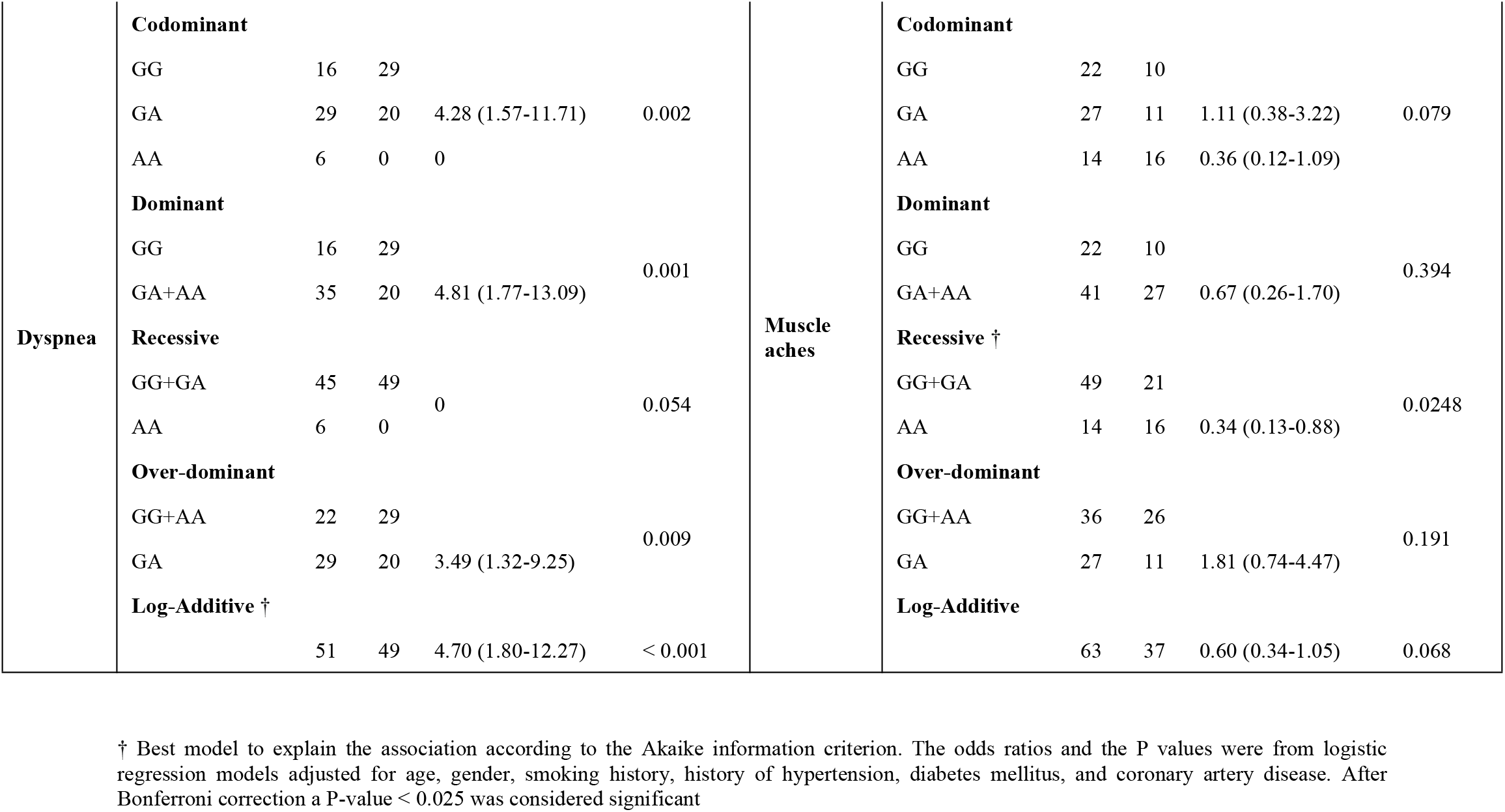
Association of *IFNAR2* rs2236757 and *OAS3* rs10735079 polymorphisms with COVID-19 signs and symptoms in all patients

| rs2236757 (*IFNAR2*) |  | Yes | No | OR (95% CI) | P-value | rs10735079 (*OAS3*) |  | Yes | No | OR (95% CI) | P-value |
| --- | --- | --- | --- | --- | --- | --- | --- | --- | --- | --- | --- |
| Sore throat | **Codominant †** |  |  |  |  | Sore throat | **Codominant** |  |  |  |  |
|  | GG | 13 | 32 |  |  |  | GG | 15 | 17 | 0.88 (0.33-2.38) |  |
|  | GA | 23 | 25 | 2.52 (1.02-6.24) | 0.011 |  | GA | 17 | 20 | 0.16 (0.04-0.58) | 0.004 |
|  | AA | 0 | 6 | 0 |  |  | AA | 4 | 26 |  |  |
|  | **Dominant** |  |  |  |  |  | **Dominant** |  |  |  |  |
|  | GG | 13 | 32 |  |  |  | GG | 15 | 17 |  |  |
|  | GA+AA | 23 | 31 | 1.98 (0.82-4.76) | 0.124 |  | GA+AA | 21 | 46 | 0.47 (0.19-1.16) | 0.099 |
|  | **Recessive** |  |  |  |  |  | **Recessive †** |  |  |  |  |
|  | GG+GA | 36 | 57 |  |  |  | GG+GA | 32 | 37 |  |  |
|  | AA | 0 | 6 | 0 | 0.026 |  | AA | 4 | 26 | 0.17 (0.05-0.55) | < 0.001 |
|  | **Over-dominant** |  |  |  |  |  | **Over-dominant** |  |  |  |  |
|  | GG+AA | 13 | 38 |  |  |  | GG+AA | 19 | 43 |  |  |
|  | GA | 23 | 25 | 2.89 (1.18-7.13) | 0.018 |  | GA | 17 | 20 | 1.86 (0.78-4.45) | 0.162 |
|  | **Log-Additive** |  |  |  |  |  | **Log-Additive** |  |  |  |  |
|  |  | 36 | 63 | 1.25 (0.61-2.57) | 0.538 |  |  | 36 | 63 | 0.44 (0.24-0.79) | 0.004 |
| Dyspnea | Codominant |  |  |  |  | Muscle aches | Codominant |  |  |  |  |
|  | GG | 16 | 29 |  |  |  | GG | 22 | 10 |  |  |
|  | GA | 29 | 20 | 4.28 (1.57-11.71) | 0.002 |  | GA | 27 | 11 | 1.11 (0.38-3.22) | 0.079 |
|  | AA | 6 | 0 | 0 |  |  | AA | 14 | 16 | 0.36 (0.12-1.09) |  |
|  | Dominant |  |  |  |  |  | Dominant |  |  |  |  |
|  | GG | 16 | 29 |  |  |  | GG | 22 | 10 |  |  |
|  | GA+AA | 35 | 20 | 4.81 (1.77-13.09) | 0.001 |  | GA+AA | 41 | 27 | 0.67 (0.26-1.70) | 0.394 |
|  | Recessive |  |  |  |  |  | Recessive † |  |  |  |  |
|  | GG+GA | 45 | 49 |  |  |  | GG+GA | 49 | 21 |  |  |
|  | AA | 6 | 0 | 0 | 0.054 |  | AA | 14 | 16 | 0.34 (0.13-0.88) | 0.0248 |
|  | Over-dominant |  |  |  |  |  | Over-dominant |  |  |  |  |
|  | GG+AA | 22 | 29 |  |  |  | GG+AA | 36 | 26 |  |  |
|  | GA | 29 | 20 | 3.49 (1.32-9.25) | 0.009 |  | GA | 27 | 11 | 1.81 (0.74-4.47) | 0.191 |
|  | Log-Additive † |  |  |  |  |  | Log-Additive |  |  |  |  |
|  |  | 51 | 49 | 4.70 (1.80-12.27) | < 0.001 |  |  | 63 | 37 | 0.60 (0.34-1.05) | 0.068 |

† Best model to explain the association according to the Akaike information criterion. The odds ratios and the P values were from logistic regression models adjusted for age, gender, smoking history, history of hypertension, diabetes mellitus, and coronary artery disease. After Bonferroni correction a P-value < 0.025 was considered significant

**Table 6:**
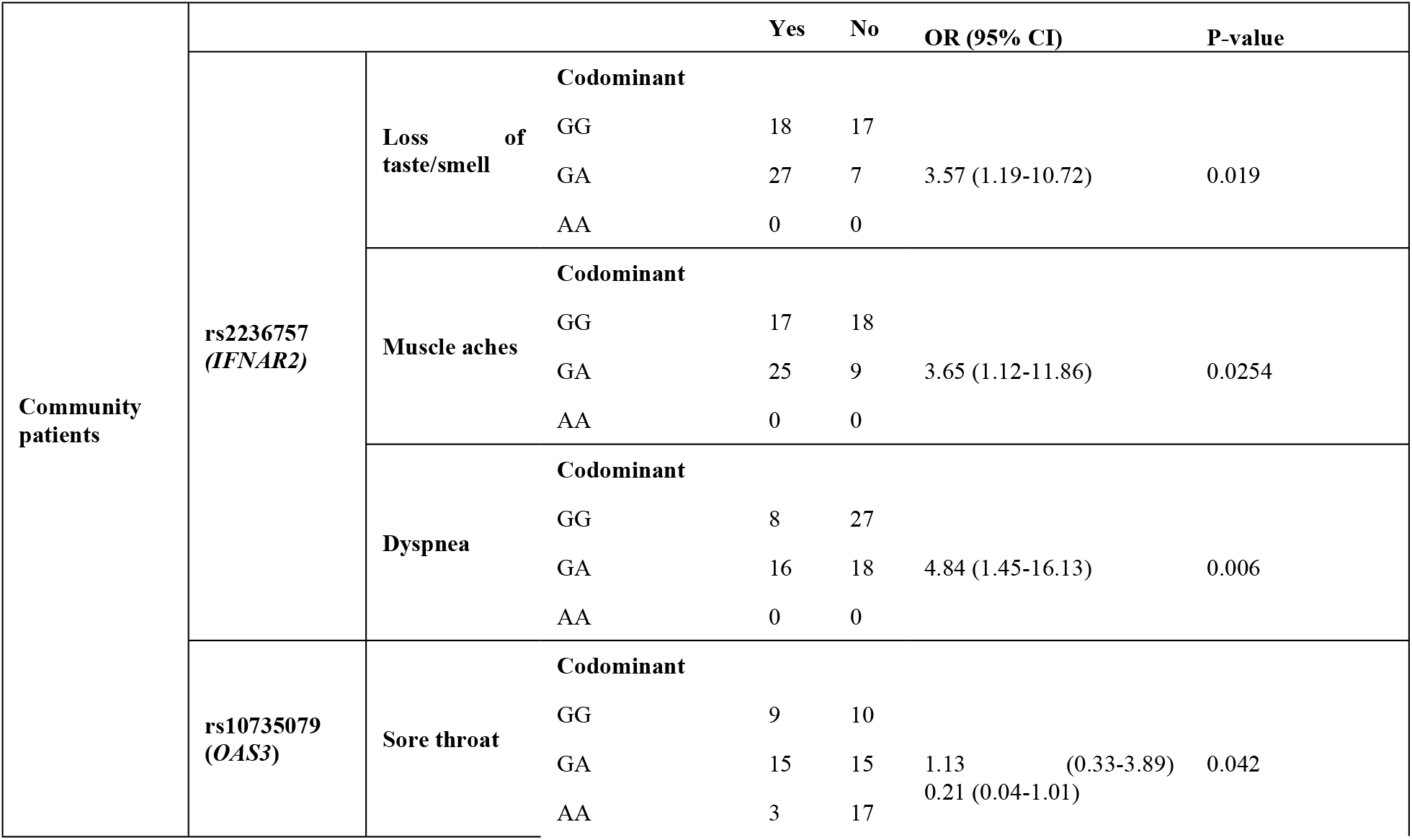
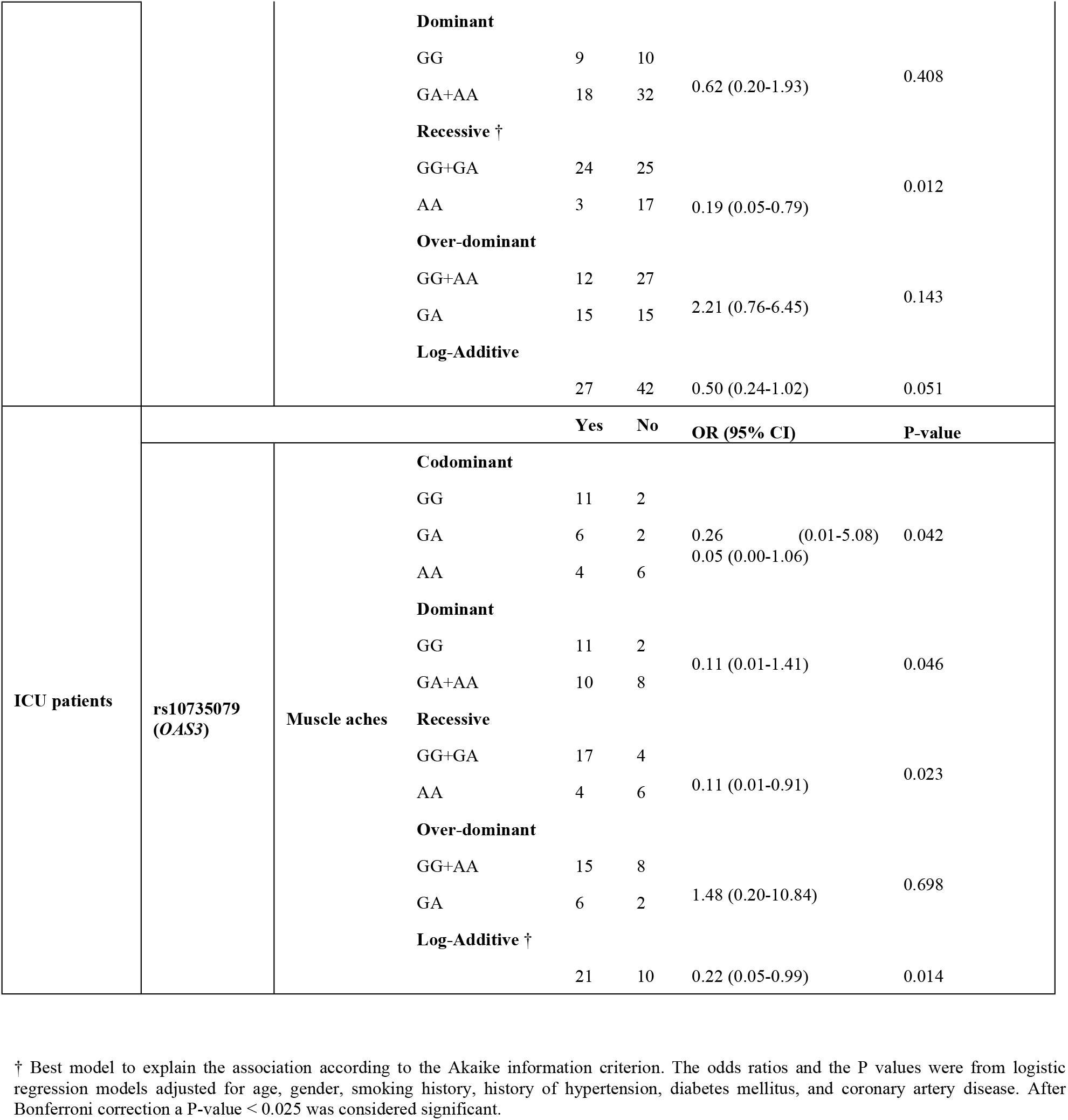
Association of *IFNAR2* rs2236757 and *OAS3* rs10735079 polymorphisms with COVID-19 signs and symptoms in the community patients and ICU patients, separately

## Discussion

COVID-19 manifestations are variable among patients, even amongst household members. Before the introduction of the COVID-19 vaccination, 33% of people with SARS-CoV-2 infection were reported to be asymptomatic [14]. Wu Z and McGoogan JM reported mild COVID-19 infection in 81% of the patients, severe disease in 14%, and critical illness in 5%[15]. Age, comorbidities, sex, and socioeconomic background play an essential role in COVID-19 severity [16–19]. Moreover, novel host genetic factors associated with COVID-19 infection and severity were identified through the collaborated community of human genetics researchers [20]. In the current study, we found that the risk allele *IFNAR2* rs2236757A has a significant association with the critical COVID-19 illness. Such an association was not present for the rs10735079 variant. The rs2236757A variant was found to be related to the critical COVID-19 illness in genome-wide significant associations. Additionally, *IFNAR2* has a causal role from a Mendelian randomization result; increased expression of the interferon receptor subunit IFNAR2 reduced the odds of severe COVID-19 (P = 0.0043) [7]. Type 1 interferons bind IFNAR2, which leads to activation and signal transduction, involving the JAK-STAT pathway [21]. Consequently, this pathway initiates antiviral activity from the target cells and induces apoptosis in infected cells [22]. The role of *IFNAR2* expression was further replicated in other Mendelian randomization studies [23–25]. OAS is a family of antiviral proteins consisting of four members, OAS1, OAS2, OAS3, and OAS-like protein [26]. Both, Interferon and virus infection stimulate the transcription of *OAS* genes in the cell [27,28]. Ribonuclease L is activated through the OAS1- to 3 proteins; products with 2’-5’ oligoadenylate synthetase activity. Ribonuclease L activation leads to the degradation of the cellular and viral RNA, resulting in the inhibition of protein synthesis and terminating viral replication [29–31]. The variant rs10735079 lies in the interferon-inducible *OAS* gene cluster (*OAS1, OAS2*, and *OAS3*) and was associated with critical COVID-19 illness (P = 1.65 × 10^8^) in genome-wide significant associations [7]. However, a similar association was not present in our study.

Different symptom clusters have a difference in in-hospital outcomes [32]. Reis et al. reported in a cross-sectional study with nearly 60,000 COVID-19 patients that fever and breathing difficulty increased the chances of hospitalization and death. However, running nose, sore throat, diarrhea, and headache were associated with lower odds of hospitalization and death; they concluded that these symptoms indicate a protective effect [33]. Sadeghifar et al. showed in their binary logistic regression model of disease outcome by disease symptoms that shortness of breath and abnormal chest radiographic findings were predictors of higher mortality. Conversely, patients with sore throats signified lower mortality [34]. Moreover, Chang et al. indicated that body temperature, chills, initial chest X-ray findings, and the presence of diabetes were significant predictors of progression to the severe stage of COVID-19 [35].

Studies that evaluate the role of human genetics in developing the different signs and symptoms of COVID-19 are scarce. Williams et al. studied 3261 same-sex twins to investigate the presence of heritable components in developing the different symptoms of COVID-19. They found heritability elements for delirium, diarrhea, fatigue, anosmia, and meal skipping [36].

Herein, we investigated the role of the rs2236757 and the rs10735079 variants in developing the different signs and symptoms of COVID-19 in all COVID-19 patients, and in the community patients and ICU patients, separately. Our results indicated that patients with the risk allele rs2236757A were more likely to have dyspnea and sore throat. Among the community patients, the risk allele rs2236757A was found to be associated with dyspnea, loss of taste or smell, and muscle aches. Surprisingly, patients with the risk allele rs10735079A were unlikely to have a sore throat or muscle aches; in particular, the community cases group was less likely to have a sore throat, and the ICU subgroup was less prone to muscle aches. Given that the risk allele rs10735079A was found to be associated with critical COVID-19 illness in multiple previous studies [7,37], the inverse association of the risk allele rs10735079A with having a sore throat might be consistent with Reis et al. and Sadeghifar et al. findings indicating that sore throat might have a protective effect against COVID-19 hospitalization and death [33,34].

It was reported that Interferon-beta inhibits SARS-CoV-2 virus replication in vitro [38]. However, clinical trials did not reveal a clear benefit from interferon therapy for hospitalized patients with severe COVID-19 [39–42]. Yet, a systematic review of five clinical trials concluded that early administration of Interferon-beta combined with other antiviral drugs is promising [43]. Low expression of *IFNAR2* has a causal role in the progression to critical COVID-19 illness [7]; our study was in line with the Pairo-Castineira et al. study which demonstrated that rs2236757A was associated with the severity of COVID-19 illness [7]. Therefore, a randomized controlled trial that examines the role of interferon therapy in COVID- 19 patients who have the rs2236757A variant or other reported variants will help to understand the role of interferon therapy and its benefits.

Our study is limited by the small number of included participants, which was in part due to the newly emerged variants of the SARS-CoV-2 virus; we could not continue recruiting patients when the new SARS-CoV-2 variants became prevalent in Palestine, as different variants may have different manifestations and pathogenesis [44,45]. Moreover, the COVID-19 vaccination was started by the government, which can alter the results. The limited number of patients in the present study may explain the lack of association between the rs10735079 polymorphism and the critical COVID-19 illness. Nonetheless, targeting families strengthen the certainty of adequate exposure to the SARS-CoV-2 virus in the control group. Additionally, the present study is the first in Palestine and one of the few studies that investigated the role of human genetic factors in COVID-19 signs and symptoms. In Conclusion, our study revealed that the *IFNAR2* rs2236757A variant is associated with critical COVID-19 illness. the risk allele rs2236757A was associated with dyspnea and sore throat while patients with the risk allele rs10735079A were unlikely to have a sore throat or muscle aches. Our study may provide preliminary results for further genetic association studies to clarify the role of human genetics in different signs and symptoms of COVID-19 and to enhance our understanding of the pathophysiology of SARS-CoV-2 infection and its complications.

## Data Availability

All relevant data are within the manuscript and its Supporting Information files.

## Acknowledgements

The authors would like to thank all the study participants

## Authors’ contributions

SE and AN designed and supervised the experiments, edited and revised the manuscript. MA analyzed the data and wrote the first draft of the manuscript. MA, OA, RA, RS, OM, TH, and ZE extracted the DNA and performed the experiments. AJ involved in patient sampling and data collection. All authors read and approved the final manuscript.

## Funding

This research received no specific grant from any funding agency.

## Availability of data and materials

Data used in this research are available from the corresponding author on request

## Declarations

### Ethics approval and consent to participate

The study procedure was approved by the research ethics committee at Al-Quds University (184/REC/2021), with implied consent from all participants.

## Competing interests

The authors declare that no competing interests exist.

## Notes

### Competing Interest Statement

The authors have declared no competing interest.

### Funding Statement

The author(s) received no specific funding for this work

